# Relative Importance of Various Inflammatory Markers and Their Critical Thresholds for COVID-19 Mortality

**DOI:** 10.1101/2021.12.24.21268371

**Authors:** Sandeep Budhiraja, Abhaya Indrayan, Poonam Das, Arun Dewan, Omender Singh, Vivek Nangia, Y P Singh, Rajesh Pandey, Ajay Kumar Gupta, Manish Gupta, Deepak Bhasin, Bhupesh Uniyal, Mohit Mathur, Mayur Pawar

## Abstract

**Background:** Various inflammatory markers are commonly assessed in many patients to help in the management of COVID-19 patients. It is not clear, though, how much risk of mortality their different levels of elevations entail, and which marker signifies more risk than others and how much. This study was undertaken to describe their levels and to answer these questions regarding eight inflammatory markers, namely, CRP, D-dimer, ferritin, IL-6, LDH, CPK, troponin-I.

**Methods:** The data were retrieved from the electronic records of 19852 CoViD-19 patients admitted to a chain of hospitals in north India from March 2020 to July 2021. Levels for most markers were available for more than 10,000 patients. In view of widely different ranges of values of different markers, we divided their values into quintiles (Qs) and studied the pattern of mortality and for running the logistic regression. In addition, logarithm transformation was also tried. The statistical distribution of the values was compared by Mann-Whitney test. Relative importance was judged by the mortality rates, area under the ROC curves (AUROCs), and the odds ratios.

**Results:** Although the mortality increased with decreasing ALC and increasing level of all the other markers, more than 70% survived even with levels in the extreme quintile. The adjusted odds ratio was the highest (7.62) for the Q5 levels of IL-6, closely followed by D-dimer (OR = 6.04). The AUROC was the highest (0.817) for LDH and the least (0.612) for CPK. However, the optimal cut-off for any marker could correctly classify not more than 80% deaths and the multivariable logistic regression could correctly classify patients with mortality in less than 24% cases.

**Conclusion:** Although elevated levels of all the markers and low values of ALC were significant risk factor but no firm evidence was available for any of the eight markers to be a major indicator of the mortality in COVID-19 unless they reach to a critical threshold. Among those studied, D-Dimer (>192 ng/mL) followed by IL-6 (>4.5 pg/mL) had stronger association with mortality even with moderate and higher end of the normal levels and LDH (>433 U/L) and troponin-I (>0.002ng/mL) with only steeply increased levels. Ferritin had modest association, and CPK, CRP and ALC were a relatively poor risk of mortality.

## INTRODUCTION

COVID-19 pandemic has already killed more than 5 million people across the world by the end of November 2021. The pandemic has continued in waves in many countries, and the disease is suspected to remain in our midst at endemic level in the long run^1^. Thus, this disease is likely to remain of clinical interest for times to come.

The disease causes cytokine storm in many patients admitted to hospitals due to exaggerated immune response. Assessment of inflammatory markers is among the commonest investigation carried out in these patients. Regular monitoring of these markers is considered to help in more effective management of the disease.

It was observed that high levels of inflammatory markers are intimately associated with increasing severity of COVID-19^2,3^ and their role in mortality has also been investigated^4,5^. However, the results with different markers are different and it is not clear what levels of these markers are helpful in assessing the risk of mortality and how they compare with one another. This study was undertaken to describe the levels of various inflammatory markers in CoViD-19 patients, to assess the association of different levels with mortality so that critical thresholds can be identified, and to investigate the relative importance of different markers as risk of mortality.

## MATERIAL and METHODS

Records of all COVID-19 patients admitted from March 2020 to July 2021 to the network of our hospitals in north India were retrieved from the electronic record system. A total of 19852 COVID-19 patients were admitted during this period. Most common age-group was 40-59 years (38.6%) and 33.6% of all patients were females.

Several inflammatory markers were investigated in these patients as ordered by the concerned clinicians, but the commonly investigated markers were C-reactive protein (CRP), D-Dimer, ferritin, interleukin-6 (IL-6), lactate dehydrogenase (LDH), creatinine phosphokinase (CPK), troponin-I, and absolute lymphocyte count (ALC). First value obtained within 5 days of admission was considered for the present analysis. Levels for more than 10,000 patients were available for most markers and the least was 4566 patients for CPK. The mortality in the patients investigated for most markers ranged from 6.04% to 9.31% against an overall mortality of 8.42%. This indicates that the markers were investigated irrespective of the severity and the available values may be a fair representation of all the admitted patients.

All the markers were evaluated by the standard method in accredited laboratories located in the respective hospitals as per the manufacturer’s manual. For CRP, latex particle immunoturbidimetric method^6^ for D-dimer, immunoturbidimetric method^7^, for ferritin, chemiluminescence method^8^, for IL-6, electrochemiluminescence method^9^, for LDH, enzymatic lactate to pyruvate method^10^, for CPK, NAC activated method^11^, for troponin-I, chemiluminescence method^12^, and for ALC, electrical impendence VCS and calculation method^13^ was used.

The shape of the distribution of the values of the markers was obtained to get a feeling of how skewed this in COVID-19 patients is. In view of widely scattered values, the levels for each marker were divided into quintiles (Qs) with each quintile comprising nearly a one-fifth of the cases for whom the level of that marker was available. Pearson correlation coefficient was obtained between the levels of these markers. Because of highly skewed distributions, Mann-Whitney test was used to compare their distribution in those who survived with those who died. The trend of mortality was studied over the quintiles. In addition, the area under the ROC curve (AUROC) for mortality was obtained. The cut-off with the highest inherent validity^14^ based on the sum of sensitivity and specificity was obtained and the mortality rate in those with less than this cut-off and more than this cut-off was obtained to check the extent of correct classification of those who survived and those who died. We also obtained the odds ratio of mortality for each marker by running multivariable logistic regression using enter method that provided adjusted odds ratio (OR) for each marker. Relatively exceedingly high mortality for the values of some markers violated the assumption of linearity for logistic regression – thus log-values were tried, and quintiles categories were used. Thus, several methods of statistical analysis were tried to find whether they give any consistent result for a valid conclusion regarding the relative importance of various markers in COVID-19 mortality and the critical threshold.

In view of the multiple testing, a *P*-value less than 0.01 was considered significant in place of the conventional 0.05. SPSS 21 was used for calculations.

## RESULTS

### Descriptive Statistics of the Levels of Different Markers

The statistical distribution of the levels of all the markers was highly skewed to the right (Figure 1) as expected and of ALC slightly skewed. The values of CRP, IL-6, and troponin-I, and, to a large extent, of ferritin followed an exponential shape with the highest number of patients (frequency) with very low levels and the frequencies showing sharp decline with increasing levels in the case of CRP, IL-6, and troponin-I, and gradual decline in the case of ferritin. Against this, the distribution pattern of D-dimer, LDH, CPK, and ALC was a typical Gamma with small frequencies at low values, steeply increasing with slightly higher values and then showing a gradual decline.

**Figure 1.**
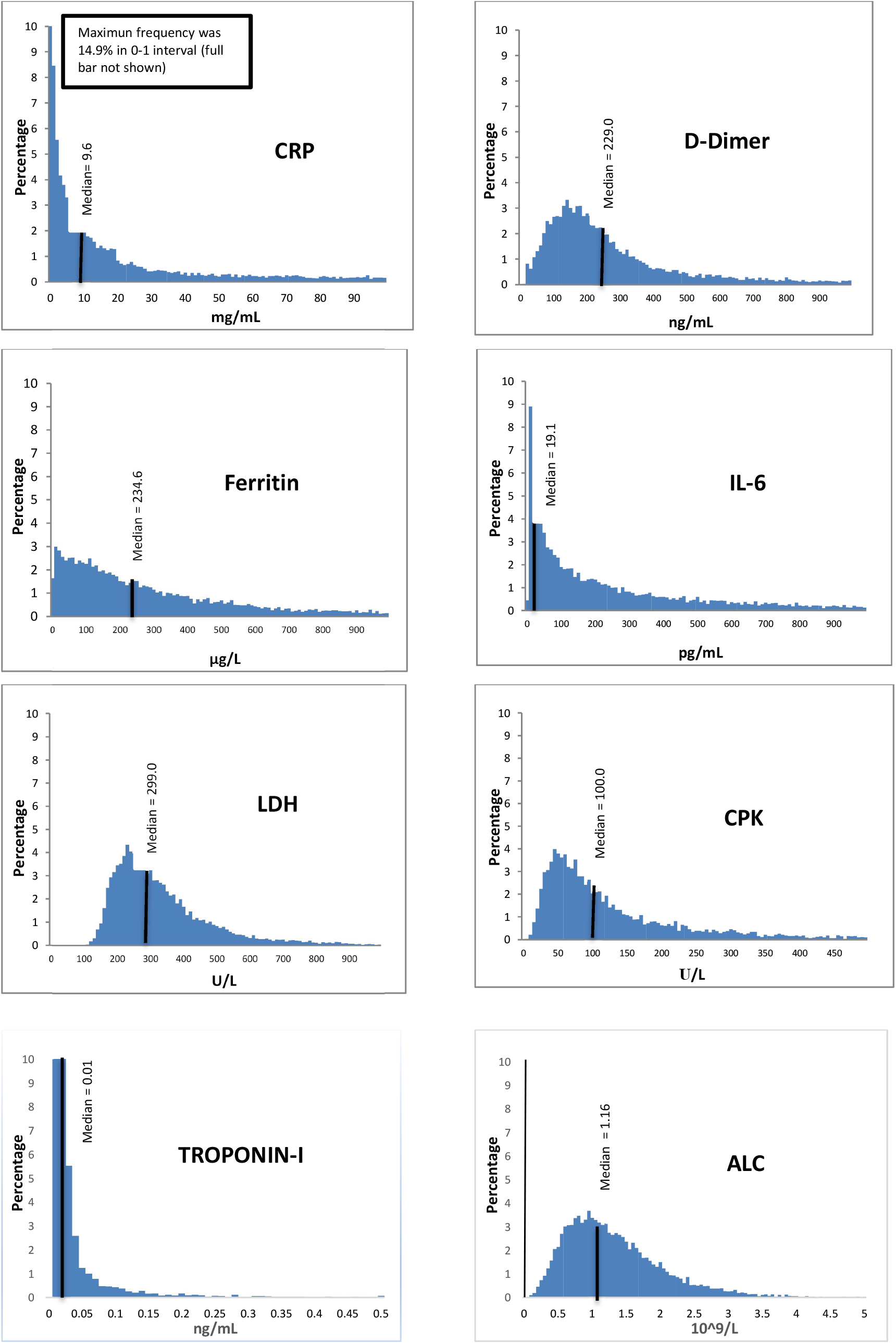
Statistical distribution of the values of different markers

In view of highly skewed distribution and huge range of levels of each of these markers, we divided the levels into 5 quintiles – each containing nearly one-fifth subjects with available levels. First 20% (Q1) mostly had levels within the normal levels and the top 20% values were exceedingly high, such as more than 53.7 mg/mL of CRP, and more than 530 ng/mL for D-dimer (Table 1). In the case of ALC, it is just the reverse.

**Table 1.** Quintiles for different markers

| Marker | Q1 | Q2 | Q3 | Q4 | Q5 |
| --- | --- | --- | --- | --- | --- |
| CRP (mg/mL) | 0-1.5 | 1.5-5.8 | 5.8-15.6 | 15.6-53.7 | >53.7 |
| D-dimer (ng/mL) | 1-127 | 127-192 | 192-278 | 278-530 | >530 |
| Ferritin (µg/L) | 2-80 | 80-172 | 172-308 | 308-580 | >580 |
| IL-6 (pg/mL) | 0-4.5 | 4.5-12.5 | 12.5-28.0 | 28.0-66.3 | >66.3 |
| LDH (U/L) | 100-220 | 220-270 | 270-332 | 332-433 | >433 |
| CPK (U/L) | 9-52 | 52-81 | 81-125 | 125-236 | >236 |
| Troponin-I (ng/mL) | 0-0.002 | 0.002-0.010 | 0.010-0.010 | 0.010-0.012 | >0.012 |
| ALC (10<sup>9</sup>/L) | 0-0.69 | 0.69-1.00 | 1.00-1.35 | 1.35-1.84 | >1.84 |

The levels with the highest frequency (mode) are shown in Table 2. Except for CRP and troponin, these frequencies are low and show highly dispersed levels of various markers in COVID-19 admitted patients.

**Table 2.** Modal intervals and the percentage of patients with low values of the markers

| Marker | n | Mode |  | Elevated* level |  | Median |
| --- | --- | --- | --- | --- | --- | --- |
|  |  | Interval | Percent in the interval | Cut-off | Percent with elevated level |  |
| CRP (mg/L) | 13247 | 0-1 | 14.9 | >3 | 70.7 | 9.6 |
| D-dimer (ng/mL) | 13095 | 140-150 | 3.3 | >250 | 44.7 | 229.0 |
| Ferritin (µg/L) | 10483 | 10-20 | 3.0 | >300 | 34.8 | 234.6 |
| IL-6 (pg/mL) | 10873 | 1-2 | 8.9 | >10 | 62.3 | 21.5 |
| LDH (U/L) | 9495 | 230-240 | 4.3 | >280 | 56.7 | 301 |
| CPK (U/L) | 4566 | 45-50 | 4.0 | >120 | 46.3 | 100.0 |
| Troponin-I (ng/mL) | 5601 | 0.01-0.02 | 11.6 | >0.04 | 64.1 | 0.01 |
| ALC (10<sup>9</sup>/L) | 16620 | 0.9-1.0 | 7.1 | <1.10 | 48.0 | 1.16 |

\* Low value in the case of ALC

Although the modes were within the usual reference range of these parameters, but more than one-half patients had elevated levels of many markers, indicating the incidence of cytokine storm in these patients. The median levels of CRP, IL-6, and ferritin were also quite high (Table 2), indicating that more than one-half COVID-19 patients had cytokine storm with respect to these markers.

Values of all the eight inflammatory markers were available for 4108 patients and the mortality in them was 8.85%. This is not much different from 8.42% in all the cases. Thus, these 4108 patients may also be a fair representation of all the cases with respect to mortality. The Pearson correlation coefficients among the levels of the inflammatory markers in these patients, though statistically significant because of large *n* in our study, were low with a maximum of 0.207 (Table 3) except 0.429 between ferritin and LDH and 0.313 between D-dimer and troponin-I levels. Thus, it looks that various markers mostly work relatively independently of one another, at least linearly because the Pearson correlation coefficient measures the strength of only the linear relationship.

**Table 3.**
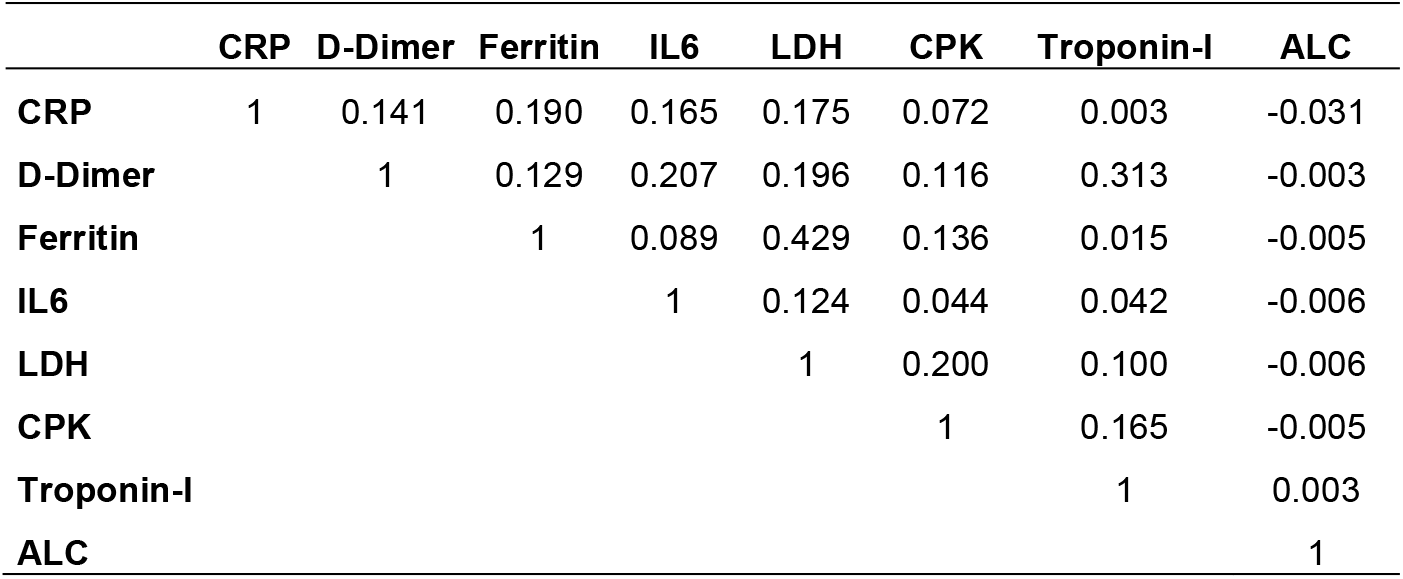
Correlation coefficients among the levels of various markers

### Levels of the Markers in the Surviving and Died Patients

The distributions of the levels of all the markers were significantly different (*P* < 0.001) in the patients who survived from those who died. The median levels of all the markers were significantly (*P* < 0.001) higher in those who died although the medians in the surviving patients were also high (low in the case of ALC) (Table 4). When the levels were divided into quintiles with nearly 20% cases in each category, the mortality showed steep rise with increasing quintiles for D-dimer and IL-6, relatively slow increase for CRP and ferritin, and not much increase in the case of CPK (Figure 2). LDH and troponin-I showed steeply high mortality risk when they reached to the top quintile (> 433 U/L for LDH and > 0.012 ng/ml for troponin-I) but not much when it is lower than this level. In the case of ALC, mortality increased as the levels declined, but the mortality was particularly high when the levels were in the first quintile (< 0.69×10^9^/L).

**Table 4.**
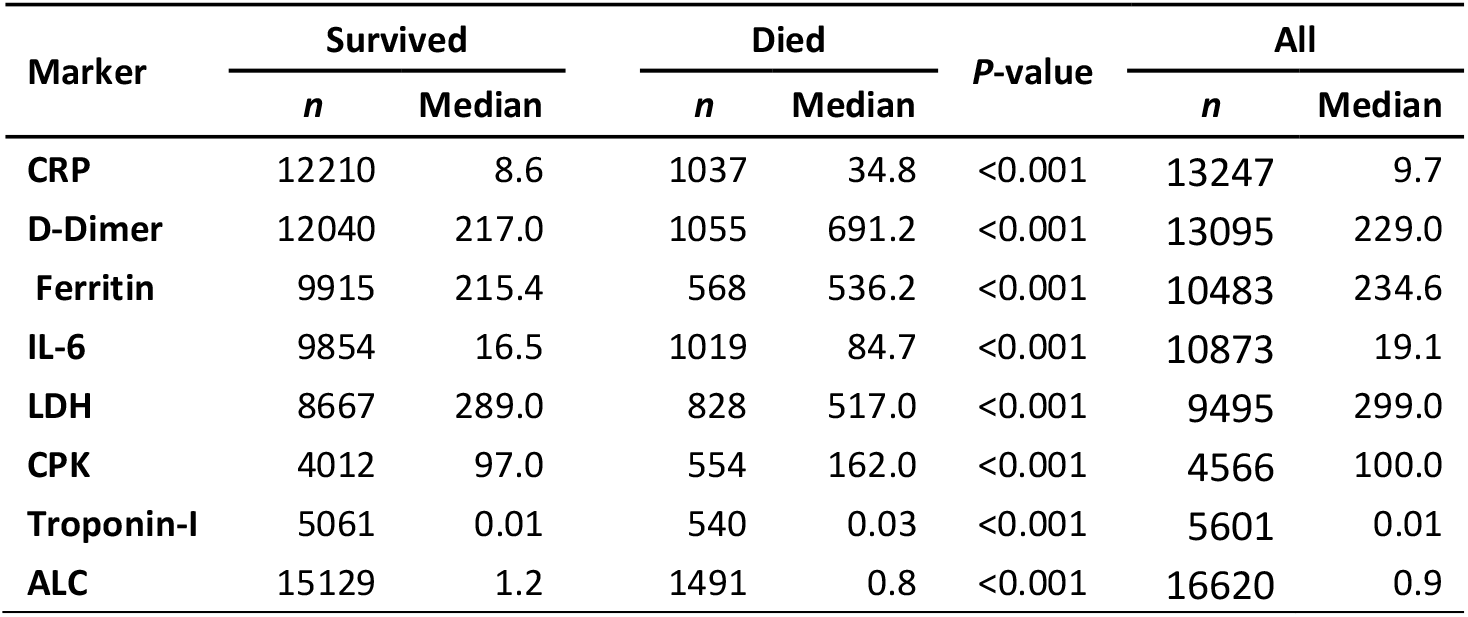
Median values of various markers in the surviving and died patients

**Figure 2.**
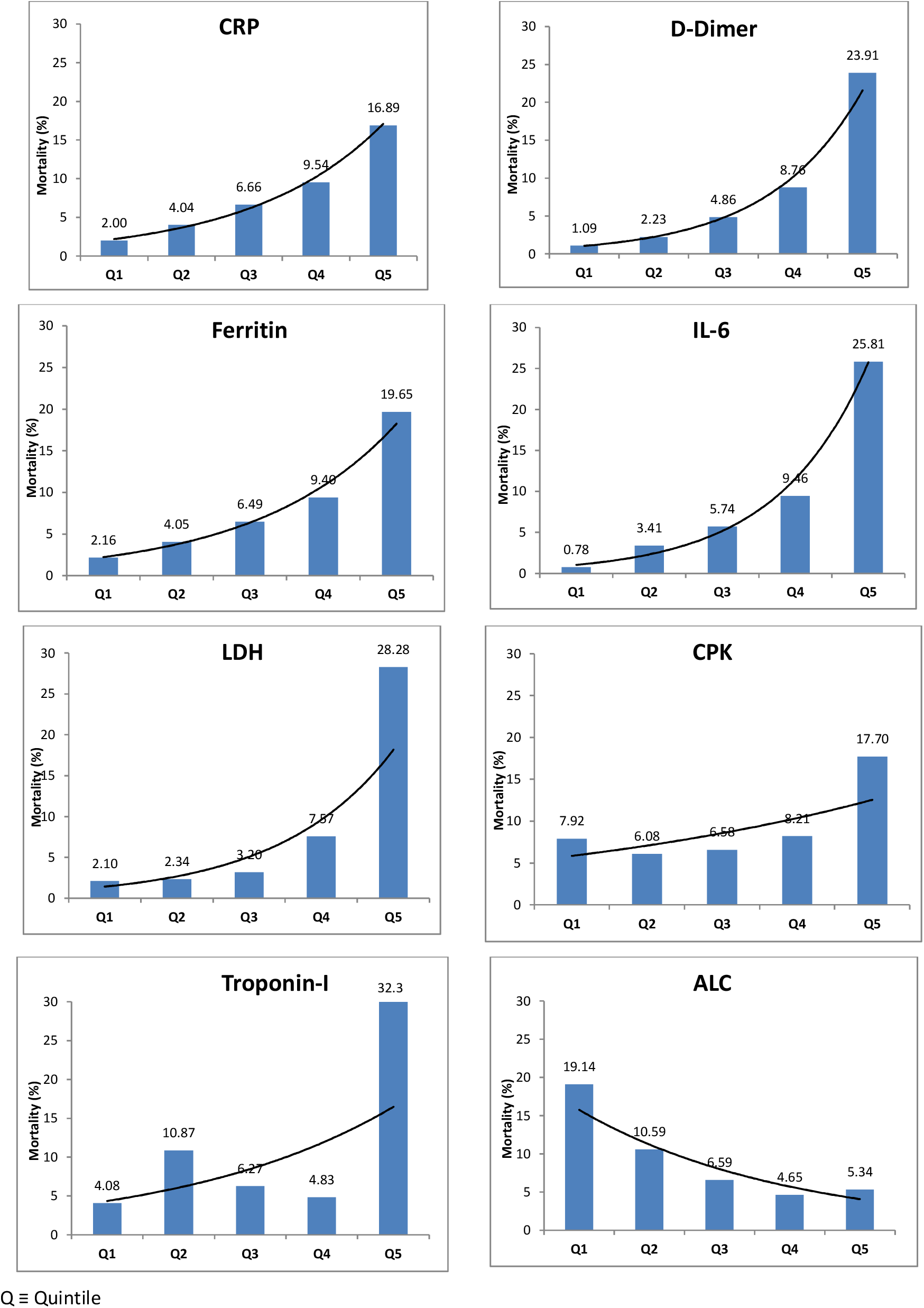
Trend of morality in different quintile categories of the markers

The mortality in patients with normal levels of various markers ranged from 1.9% to 10.8% which rose to 10.7% to 15.1% in those with elevated (low in the case of ALC) levels, and 38.0% in the case of troponin (Table 5). The highest ratio of mortality (nearly 1:8) was with elevated levels of troponin followed by D-dimer (nearly 1:7). The relative mortality in those with raised CPK levels was only one-and-a half times of the mortality in those with normal levels.

**Table 5.** Mortality in patients with normal levels and elevated levels

| Marker | Normal levels* |  |  | Elevated** levels |  |  | Mortality ratio |
| --- | --- | --- | --- | --- | --- | --- | --- |
|  | n | Deaths | Percent Mortality | n | Deaths | Percent Mortality |  |
| CRP | 4888 | 143 | 2.9 | 8359 | 894 | 10.7 | 3.7 |
| D-Dimer | 6875 | 134 | 2.0 | 6220 | 921 | 14.8 | 7.6 |
| Ferritin | 6458 | 125 | 1.9 | 4025 | 475 | 11.8 | 6.1 |
| IL-6 | 2990 | 65 | 2.2 | 7883 | 954 | 12.1 | 5.6 |
| LDH | 2953 | 56 | 1.9 | 6542 | 772 | 11.8 | 6.2 |
| CPK | 3160 | 342 | 10.8 | 1406 | 212 | 15.1 | 1.4 |
| Troponin-I | 4772 | 225 | 4.7 | 829 | 315 | 38.0 | 8.1 |
| ALC | 9955 | 491 | 4.9 | 6665 | 1000 | 15.0 | 3.0 |

\* At the cut-off given in Table 2
\*\* Low levels in the case of ALC

### Area Under the ROC Curves

The ROC curves are plots of true positivity rate against false positivity rate where positivity here refers to mortality. The AUROC, which indicates the efficacy of the levels for identifying survival and mortality, was the highest (0.817) for LDH, followed by troponin-I, D-Dimer, and IL-6 with AUROC = 0.807, 0.797, and 0.793, respectively (Figure 3). The least was 0.612 for CPK.

**Figure 3.**
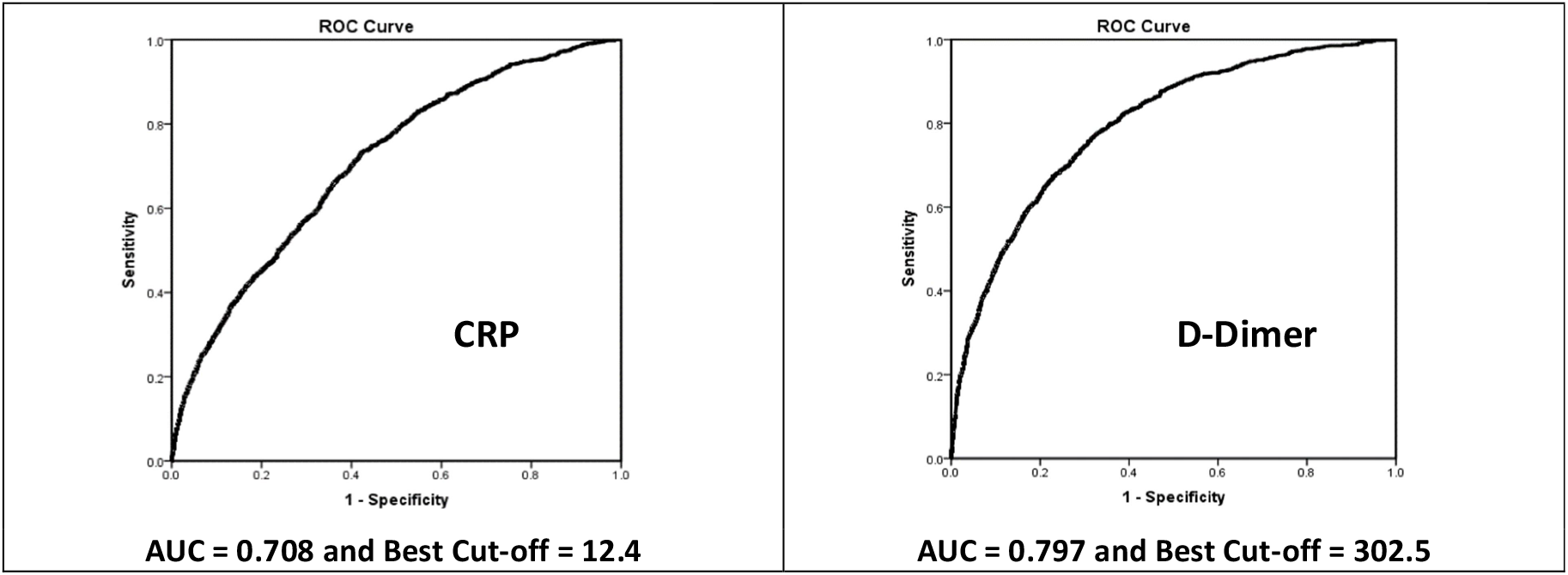
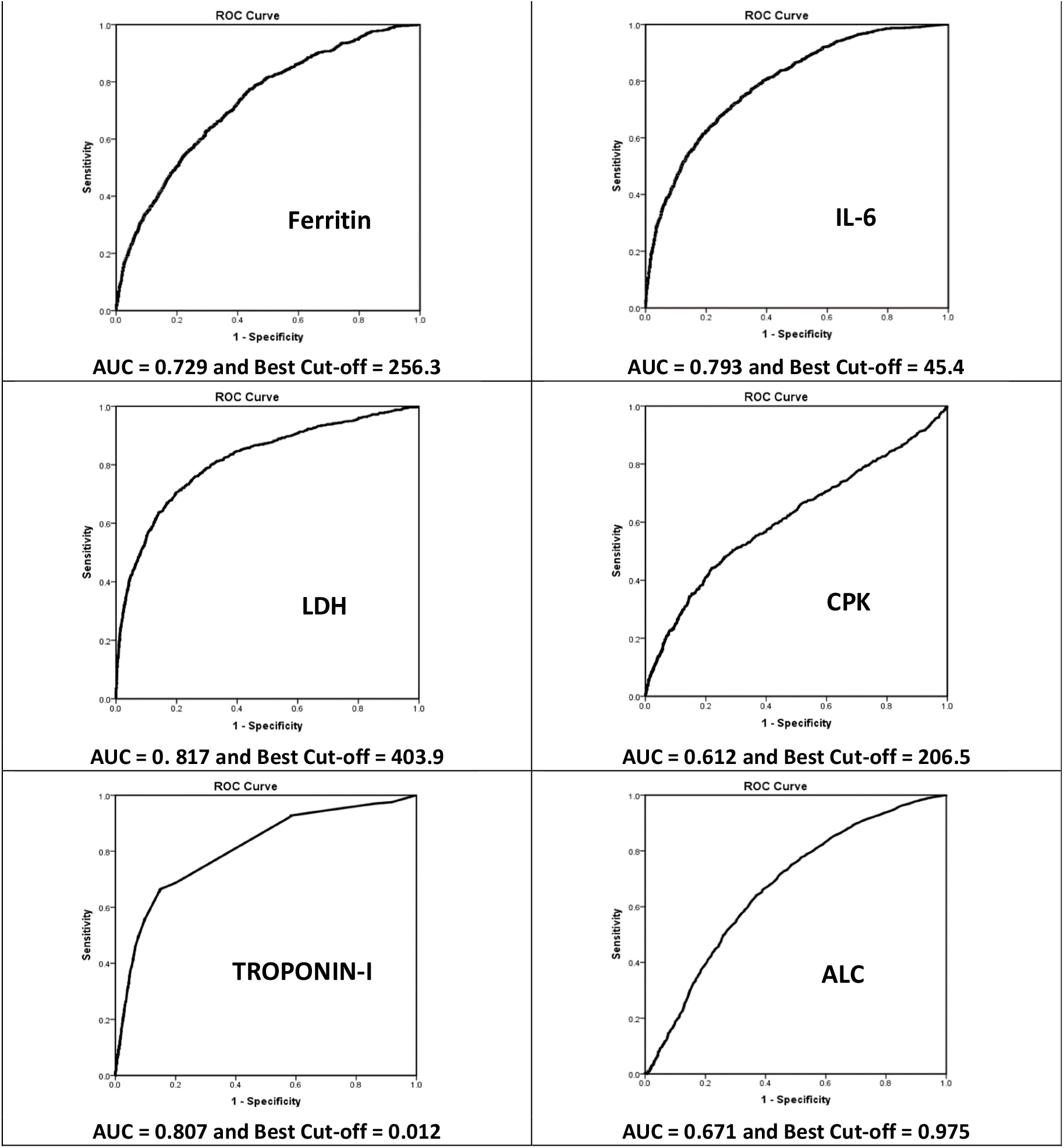
ROC curves for different markers

The most correct classification of mortality was by D-dimer level of more than 302.5 ng/mL but that too did not exceed 80% (Table 6). The correct classification of survivals by the levels less than these cut-offs was even lower at nearly 70%. The survival was best detected (88.2%) by CPK values less than 206.5 μg/L but this cut-off correctly classified mortality in only 44.0% cases (Table 6). Considering correct classification of both deaths and survivals together, the best overall accuracy was 82.8% by CPK with the optimal cut-off 206.5 μg/L, followed by IL-6 with cut-off 45.4 pg/mL. LDH with cut-off 403.9 U/L also had overall accuracy of 82.0%. No marker was able to correctly classify deaths and survivals in more than 83% cases, and most of the correct classifications were for survivals and not many for mortality. Other markers had lower performance. The overall accuracy was least (61.6%) by CRP at the optimal cut-off of 12.4 mg/L.

**Table 6.** Correct classification of deaths and survival by the “Optimal” cut-off

| Marker | Total cases | Optimal cut-off* | Deaths |  |  | Discharged |  |  | Overall accuracy (%) |
| --- | --- | --- | --- | --- | --- | --- | --- | --- | --- |
|  |  |  | Total deaths | Deaths in those with values higher than the optimal cut-off** | Correctly classified for mortality (%) | Total survived | Survive in those with values lower than the optimal cut-off | Correctly classified for survival (%) |  |
| CRP | 13247 | 12.4 | 1037 | 760 | 73.3 | 12210 | 7337 | 60.1 | 61.1 |
| D-Dimer | 13095 | 302.5 | 1055 | 840 | 79.6 | 12040 | 8433 | 70.0 | 70.8 |
| Ferritin | 10483 | 256.3 | 568 | 749 | 61.4 | 9915 | 6167 | 62.2 | 62.2 |
| IL-6 | 10873 | 45.3 | 1019 | 706 | 69.3 | 9854 | 8290 | 84.1 | 82.7 |
| LDH | 9495 | 403.9 | 828 | 582 | 70.3 | 8667 | 7206 | 83.1 | 82.0 |
| CPK | 4566 | 206.5 | 554 | 244 | 44.0 | 4012 | 3537 | 88.2 | 82.8 |
| Troponin-I | 5601 | 0.012 | 540 | 340 | 63.0 | 5061 | 4140 | 81.8 | 80.0 |
| ALC | 16620 | 0.975 | 1491 | 941 | 63.1 | 15129 | 9299 | 61.5 | 61.6 |

\*Best cut-off for classifying survival and death (highest sensitivity + specificity)
\*\* Lower values in the case of ALC

Extremely high levels of these markers were also seen, though rare, and turned out to be associated with high mortality. There were 130 (1.4%) cases with LDH level 1000 U/L or more and 93 (71.5%) of them died. Similarly, 113 (0.9%) cases had D-dimer level 10,000 ng/mL or more and 59(52.2%) died, 97 (0.8%) cases with IL-6 level at least 1,000 pg/mL and 62(63.9%) died. Only 4 (0.1%) cases had ferritin level 10,000 μg/L or more and 2(50.0%) died. Thus, extremely high levels of these markers had high mortality.

### Logistic Regression

Steeply increasing mortality with increasing levels of some markers violates the assumption of linearity for valid logistic regression results when the levels are considered as the continuous variables. Trend in Figure 2 is a testimony for this. We tried log transformation (Supplement) but that too did not help because the mortality was exceedingly high for some levels (Figure S1). As a remedy, we used quintile categories for each marker without considering the ingredient. This obviates the need for linearity^15^. The results of multivariable logistic regression are in Table 7. Now the highest odds ratio (aOR) with Q1 as the reference category, adjusted for other markers, was 8.40 for Q2 levels of troponin-I, which could be an aberration because there was no trend. For stable trend, highest aOR was 7.62 for Q5 of IL-6 followed by 6.04 for Q5 levels of D-dimer and the lowest was 0.81 for Q4 of CPK though not statistically significant (*P* = 0.326). These ORs indicate that the risk of mortality increased to as much as 7 times when the levels of IL-6 and D-dimer reached to the top quintile. At the stricter 1% level of significance, CRP, ferritin, CPK, and ALC did not provide statistically significant aOR for any quintile category. Q5 (> 433 U/L) was significant for LDH, and Q3, Q4 and Q5 for D-Dimer (>192 ng/mL) and all quintiles after the first for IL-6 (> 4.5 pg/mL). Except for Q4, troponin-I levels more than 0.002 ng/mL also had significant aOR for mortality. The multivariable logistic regression, which considered all the markers together for both mortality and survival, could correctly classify 91.7% cases in all, including survivals, but only 23.2% deaths were correctly classified. This indicates minor association of these markers with overall deaths and indicates that other factors also contributed to the mortality.

**Table 7.** Results of multivariable logistic regression for mortality

| Marker | Quintile | P-value | aOR | 95% CI for aOR |  |
| --- | --- | --- | --- | --- | --- |
|  |  |  |  | Lower | Upper |
| CRP | Q1 |  | Reference |  |  |
|  | Q2 | 0.539 | 1.23 | 0.64 | 2.37 |
|  | Q3 | 0.264 | 1.42 | 0.77 | 2.65 |
|  | Q4 | 0.483 | 1.24 | 0.68 | 2.24 |
|  | Q5 | 0.317 | 1.34 | 0.76 | 2.37 |
| D-Dimer | Q1 |  | Reference |  |  |
|  | Q2 | 0.205 | 1.60 | 0.77 | 3.32 |
|  | Q3 | 0.002 | 2.90 | 1.47 | 5.70 |
|  | Q4 | 0.001 | 3.25 | 1.66 | 6.35 |
|  | Q5 | <0.001 | 6.04 | 3.09 | 11.79 |
| Ferritin | Q1 |  | Reference |  |  |
|  | Q2 | 0.575 | 1.19 | 0.65 | 2.16 |
|  | Q3 | 0.442 | 1.25 | 0.71 | 2.21 |
|  | Q4 | 0.685 | 1.13 | 0.63 | 2.00 |
|  | Q5 | 0.154 | 1.50 | 0.86 | 2.61 |
| IL-6 | Q1 |  | Reference |  |  |
|  | Q2 | 0.003 | 3.53 | 1.52 | 8.18 |
|  | Q3 | 0.001 | 3.91 | 1.72 | 8.93 |
|  | Q4 | 0.002 | 3.60 | 1.58 | 8.18 |
|  | Q5 | <0.001 | 7.62 | 3.41 | 17.04 |
| LDH | Q1 |  | Reference |  |  |
|  | Q2 | 0.892 | 1.04 | 0.56 | 1.97 |
|  | Q3 | 0.652 | 0.87 | 0.47 | 1.61 |
|  | Q4 | 0.607 | 1.17 | 0.65 | 2.11 |
|  | Q5 | <0.001 | 2.90 | 1.63 | 5.16 |
| CPK | Q1 |  | Reference |  |  |
|  | Q2 | 0.599 | 0.89 | 0.56 | 1.39 |
|  | Q3 | 0.573 | 0.88 | 0.57 | 1.36 |
|  | Q4 | 0.326 | 0.81 | 0.53 | 1.23 |
|  | Q5 | 0.701 | 1.08 | 0.73 | 1.59 |
| Troponin-I | Q1 |  | Reference |  |  |
|  | Q2 | 0.008 | 8.40 | 1.75 | 40.27 |
|  | Q3 | <0.001 | 2.23 | 1.47 | 3.37 |
|  | Q4 | 0.232 | 1.56 | 0.75 | 3.26 |
|  | Q5 | <0.001 | 5.11 | 3.36 | 7.76 |
| ALC | Q1 |  | Reference |  |  |
|  | Q2 | 0.018 | 1.70 | 1.09 | 2.64 |
|  | Q3 | 0.075 | 1.51 | 0.96 | 2.38 |
|  | Q4 | 0.994 | 1.00 | 0.60 | 1.66 |
|  | Q5 | 0.836 | 0.94 | 0.55 | 1.62 |

aOR: adjusted odds ratio

## DISCUSSION

Several meta-analyses^2, 4, 5, 16, 17^ and individual studies^18, 19^ have concluded that elevated levels of inflammatory markers are associated with severity and mortality in COVID-19 cases. This anyway is obvious because the elevated (low of ALC) levels of these markers indicate greater tissue damage that leads to deterioration of the condition of the patient. We could not locate any specific information in the literature on the relative importance of various markers as a risk factor for mortality and on the critical threshold. Also, all the studies so far, including meta-analyses, have relatively small sample whereas we have analysed values of more than 10,000 patients for 5 of the 8 markers and a substantially large minimum sample of 4566 for CPK. Thus, our results are likely to have better reliability.

We could not locate any article that describes the level of inflammatory markers in COVID-19 patients in such a detail. A typical Gamma shape of the distribution of values of D-dimer, LDH, CPK, and ALC, and its special case exponential distribution of the values of CRP, ferritin, IL-6, and troponin could help anticipate the pattern of their level in future COVID-19 patients and to apply the right statistical method for their analysis when an exact parametric analysis is required.

Quantile values presented by us for admitted COVID-19 patients describe, among others, the bottom 20% values and the top 20% values (Table 1). These were not reported earlier. Elevated values of D-dimer, ferritin, CPK, and troponin-I, and low values of ALC were seen in less than one-half of the cases but CRP, IL-6, and LDH were elevated in 60-70 percent cases (Table 2). The median, for example 19.1 pg/mL for IL-6, tells that one half of the admitted patients had level less than this value and the other half the higher levels. An interesting finding from our analysis is low correlation between the values of the most markers (Table 3), indicating they generally operate independently of one another. However, the correlation between ferritin and LDH was 0.427, which is an exception. Similarly, there was a correlation 0.313 between D-dimer and troponin. Such a correlation between these two markers has also been observed in acute myocardial infarction^20^.

Our major concern in this communication is to investigate the relative importance of various inflammatory markers in mortality and to try to find the critical thresholds. These markers are known as sensitive indicators of severity, but no work seems to have been done on these aspects. The elevated (low of ALC) level of these markers in the early phase of the disease can alert the clinicians regarding the possibility of deterioration of the patient, and their relative importance can help in better and more evidence-based clinical decision-making process.

In a systematic review of 28 studies (n = 4663), Zhang et al.^16^ observed increased CRP in 73.6% patients, increased IL-6 in 53.1%, and increased LDH in 46.2%. We observed elevated LDH in more (56.7% - Table 2) cases but elevated CRP and IL-6 in nearly the same percentage of cases as reported by them. The meta-analysis of 7 studies in the same paper by Zhang et al. found increased CRP and increased LDH significantly associated with severity. They studied other markers also but not the ones we analyse in this report.

The meta-analysis of 56 studies (n = 8719) by Ji et al.^17^ reported weighted mean difference of CRP, IL-6, and other markers between severe and non-severe cases, and died and surviving patients. They concluded that the severity is associated with higher levels of inflammatory markers. In a review of 72 studies, Tjendra et al.^5^ studied multiple markers and reported that ‘markedly’ elevated levels were associated with the severity of disease. Loomba et al.^4^ analysed 10 studies (n = 1584) and reported significant differences in the levels of various markers between the patients who survived and who did not^2^ included 23 studies (n = 4313) in their meta-analysis and found significantly higher levels of CRP and IL-6, among others, in severe patients.

Among individual studies, Henry et al.^18^, in a pooled analysis of 9 studies (n = 1532), reported elevated LDH levels associated with 6-fold increase in odds of developing severe disease and 16-fold increase in odds of mortality. Their report suggests that, on average, nearly 90% of severe patients had elevated LDH against only nearly 32% in non-severe patients. In a study of 923 patients in China, Zhang et al.^19^ concluded that ALC levels remaining low after the first week following symptoms onset are highly predictive of in-hospital death.

In an IL-6-based mortality risk model^21^ on patients in Spain found IL-6, CRP, LDH, ferritin, and D-dimer had AUROC > 0.70 and described them as ‘predictive’ of mortality. The study by Marimuthu et al.^21^ in India reported highest AUROC = 0.740 for IL-6 among the five markers they studied. These studies are similar to ours but were based only on 611 and 221 patients, respectively. The threshold of 0.70 for AUROC chosen by them looked too low to us for ‘prediction’ of mortality. We also observed AUROC > 0.70 for all the markers except CPK and ALC, but the AUROC must be at least 0.80 to have ‘good’ inherent validity^14^. With this criterion, we found only LDH and troponin as a good indicator of mortality with AUROC exceeding 0.80. D-dimer and IL-6 were close with AUROC 0.797 and 0.793, respectively.

The optimal cut-off revealed by the model proposed by Laguna-Goya et al.^21^ had 88% sensitivity and 89% specificity for mortality which is good, but they have not provided details of how these were obtained. Marimuthu et al.^22^ reported that IL-6 level > 60.5 pg/mL and D-dimer level > 0.5 μg/mL ‘predicted’ in-hospital mortality with sensitivity 80% and 76.7%, respectively. However, the specificity was low at 65% and 60%, respectively.

Our quintiles-based multivariable logistic model could correctly classify only 23.2% deaths but was excellent with 98.4% correct classification of survivals. Low coverage of mortality may have happened because this model considered all the markers together with no fixed threshold and optimized the classification of both the deaths and survivals.

All the studies, including ours, overwhelmingly suggest that the elevated (low of ALC) levels of the inflammatory markers in COVID-19 patients are significantly associated with death. To us this is obvious. No study seems to have commented on which marker is more important than others as a risk of mortality and what are the critical thresholds. We have done three types of analyses to investigate the association of different levels with mortality. One, mortality at the conventional normal cut-offs. Two, to find the optimal cut-offs that correctly classify both survival and mortality simultaneously. Three, to identify the critical thresholds with a high risk of mortality.

Use of the term ‘predictivity’ by some authors^18,19,21^ seems misplaced because the studies were based on records where deaths already occurred with several factors playing their role, and not prospective studies to find the mortality at different levels of the markers.

Correct classification of deaths and survival in a retrospective study cannot be interpreted as predictive. Predictivity has causal overtones whereas the study is for association only. Thus, we avoid the term predictivity and discuss only the association that could at best be called a significant risk factor. In our case, even a significant association does not necessarily translate into the significant risk of mortality because we have a huge sample size that makes it easy to obtain statistical significance even at strict level of 1%.

The ROC analyses show that the best overall accuracy for correct classification of mortality and survival was achieved by CPK at the optimal cut-off of 206.5 U/L but that too did not exceed 83% (Table 6). This also substantially included correct survivals but misclassified many deaths. Thus, the marker that can reliably identify the mortality as well as survival is illusive. Among those studied, our results suggest that the levels of IL-6, CPK, LDH, CPK, and troponin-I have relatively more importance for correctly classifying both mortality and survival. The performance of D-dimer was middling and the role of CRP, ferritin, and ALC was relatively poor. Quintile analysis (Figure 2) and the logistic results (Table 7) show that among the individual markers, the risk of mortality steeply increased with the levels of IL-6 and D-dimer, and not so much with the elevated levels of other markers we studied. LDH became a substantial risk when it reached its highest quintile (> 433 U/L) but troponin, IL-6, and D-dimer started becoming higher risk at a relatively lower level (> 0.012 ng/mL, 4.5 pg/mL, and 192 ng/mL, respectively). These include values at the upper end of the conventional normal levels. These are the thresholds that require special attention in COVID-19 patients. CPK, CRP and ALC seemed to have a poor association with mortality and ferritin expressed modest risk.

## CONCLUSION

Our results suggest that increase in IL-6 and D-dimer should be tracked more carefully in COVID-19 patients, troponin-I and LDH at a high level, increasing ferritin level carries a modest risk, and CPK, CRP, and ALC are not of much significance for mortality.

## Data Availability

All data produced are available online at EHR

## Contributions of authors

SB designed the study concept, finalized the draft, and contributed patients for the study, AI designed the study concept, guided the statistical analysis, and contributed to the draft, MP did the statistical analysis and prepared the graphs, PD provided inputs on lab data and interpretation, the remaining are clinicians (Critical Care) contributed patients/laboratory data and/or treated patients included in the present study.

## Ethics Committee Approval

The study was approved by the Institutional Ethics Committee, Max Super Speciality Hospital (A unit of Devki Devi Foundation), Address: Service Floor, Office of Ethics Committee, East Block, next to Conference Room, Max Super Speciality Hospital, Saket (A unit of Devki Devi Foundation), 2, Press Enclave Road, Saket, New Delhi – 110017 vide ref. no. BHR/RS/MSSH/DDF/SKT-2/IEC/IM/21-32 dated 23^rd^ December 2021. The IEC provided no objection and approved the publication of this manuscript.

## Consent

All the admitted patients gave a prior consent for their anonymised data to be used for research purposes.

## Supplement

Figure S1 shows that the trend of mortality with increasing values of each marker is not linear even when the values are on log-scale. For completeness, the odds ratios obtained with the log-values are given in Table S1 but these are not valid because of violation of the linearity assumption. Thus, we used quintiles as categories for running the logistic regression and not as continuous variables. The results are in Table 7 of the text.

**Figure S1.**
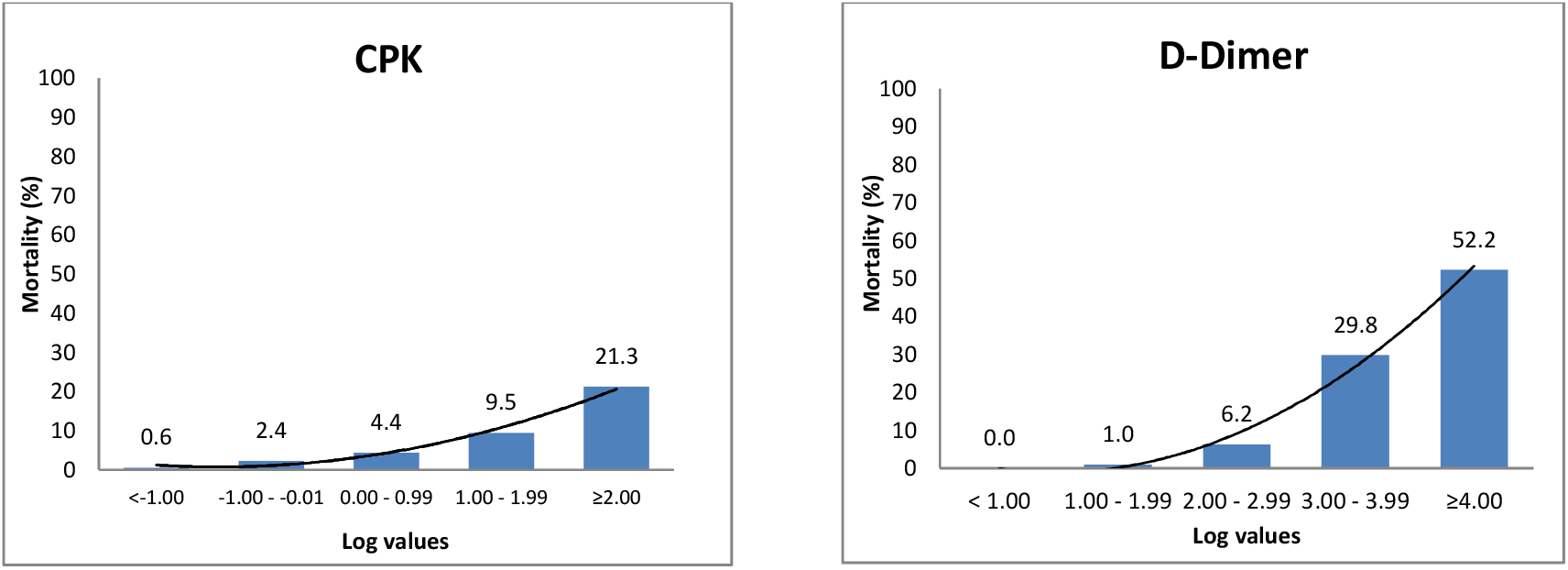
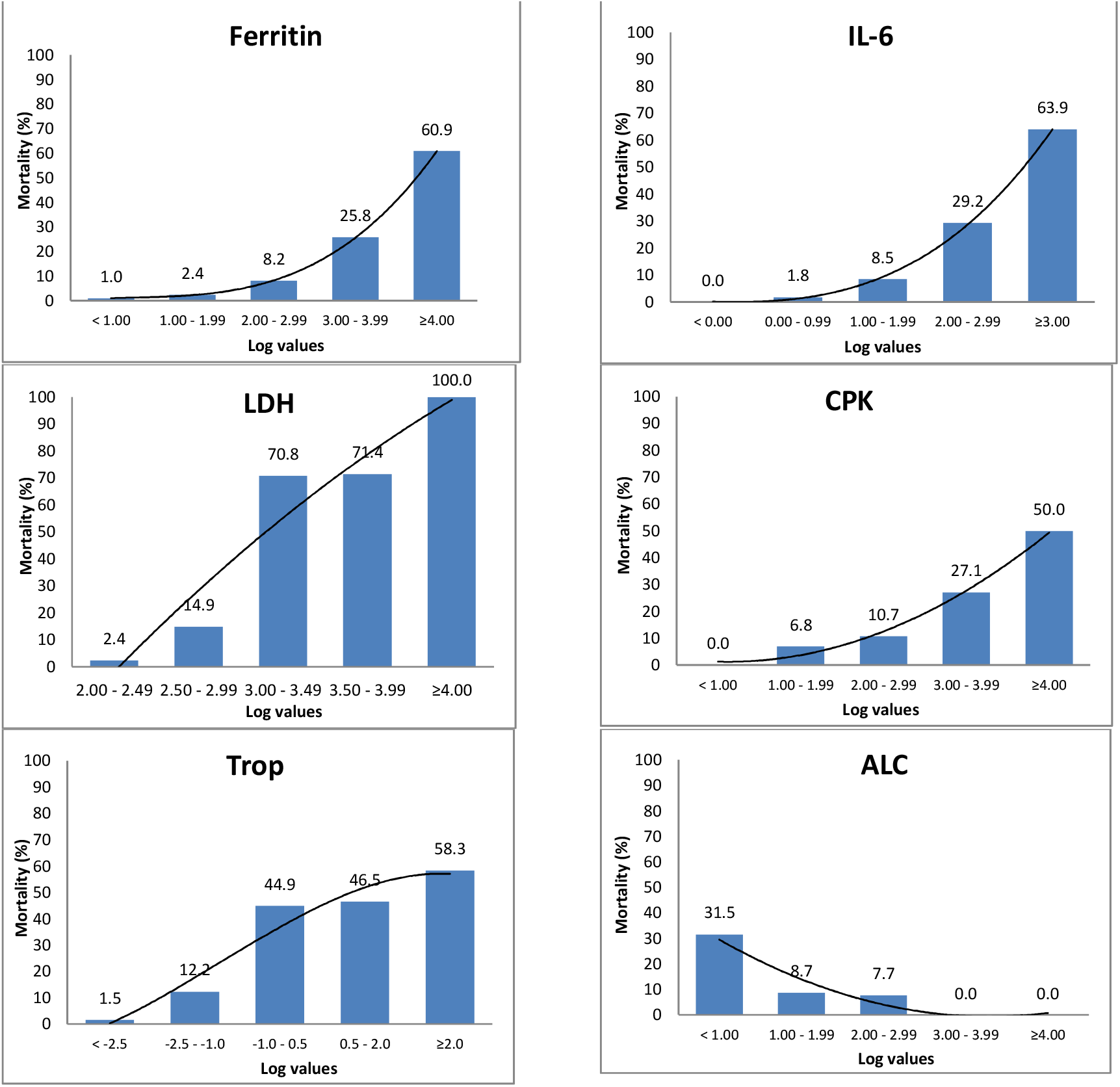
Trend of mortality with log values of different markers

**Table S1.** Multivariable logistic regression with log transformation of the levels of the markers

| Marker | P-value | aOR | 95% CI for aOR |  |
| --- | --- | --- | --- | --- |
|  |  |  | Lower | Upper |
| Log10_CRP | 0.168 | 1.13 | 0.95 | 1.35 |
| Log10_Dimer | <0.001 | 2.46 | 1.97 | 3.07 |
| Log10_Ferritin | 0.012 | 1.36 | 1.07 | 1.72 |
| Log10_IL6 | <0.001 | 1.95 | 1.60 | 2.37 |
| Log10_LDH | <0.001 | 2.30 | 1.67 | 3.15 |
| Log10_CPK | 0.101 | 1.22 | 0.96 | 1.56 |
| Log10_Trop | <0.001 | 2.29 | 1.95 | 2.70 |
| Log10_ALC | 0.001 | 0.48 | 0.31 | 0.74 |

